# SARS-CoV-2 molecular testing and whole genome sequencing following RNA recovery from used BinaxNOW COVID-19 Antigen Self Tests

**DOI:** 10.1101/2023.01.09.23284337

**Authors:** Phuong-Vi Nguyen, Ludy Registre Carmola, Ethan Wang, Leda Bassit, Anuradha Rao, Morgan Greenleaf, Julie A. Sullivan, Greg S. Martin, Wilbur A. Lam, Jesse J. Waggoner, Anne Piantadosi

**Affiliations:** Emory University Department of Medicine, Atlanta, GA, USA; Atlanta Center for Microsystems-Engineered Point-of-Care Technologies, Atlanta, GA, USA; Emory University Department of Pathology and Laboratory Medicine, Atlanta, GA, USA; Laboratory of Biochemical Pharmacology, Emory University, Atlanta, GA, USA; Emory University Department of Pediatrics, Atlanta, GA, USA; Wallace H. Coulter Department of Biomedical Engineering, Georgia Institute of Technology and Emory University, Atlanta, GA, USA

**Keywords:** SARS-CoV-2, sequencing, COVID-19, rapid test, RT-PCR

## Abstract

Widespread use of over-the-counter rapid diagnostic tests for SARS-CoV-2 has led to a decrease in availability of clinical samples for viral genomic surveillance. As an alternative sample source, we evaluated RNA isolated from BinaxNOW swabs stored at ambient temperature for SARS-CoV-2 rRT-PCR and full viral genome sequencing. 81 of 103 samples (78.6%) yielded detectable RNA, and 46 of 57 samples (80.7 %) yielded complete genome sequences. Our results illustrate that SARS-CoV-2 RNA extracted from used Binax test swabs provides an important opportunity for improving SARS-CoV-2 genomic surveillance, evaluating transmission clusters, and monitoring within-patient evolution.

## Introduction

Throughout the COVID-19 pandemic, phylogenetic studies of severe acute respiratory syndrome coronavirus 2 (SARS-CoV-2) have been instrumental in monitoring the emergence and spread of new variants, confirming diagnostic test performance, and estimating vaccine and anti-viral treatment efficacy (1-4). Monitoring is typically achieved by whole genome sequencing, but real-time reverse transcription PCRs (rRT-PCRs) have also been employed to provide rapid genotyping and assist with sample selection for sequencing workflows (5-8). Current SARS-CoV-2 genomic surveillance approaches rely upon residual clinical samples from multiple sources affiliated with the medical establishment, such as clinics, drive-through testing sites, and hospitals.

Corresponding with the rise in omicron variant transmission in the United States from December 2021 to early 2022, there was a significant increase in at-home testing for SARS-CoV-2 (9). Over-the-counter tests provide rapid and convenient testing. However, results are rarely reported to public health authorities, and samples are lost to current genomic surveillance approaches (9, 10). The BinaxNOW COVID-19 Antigen Self Test (hereinafter referred to as the Binax test; Abbott, Chicago, IL) is a common over-the-counter method that provides qualitative detection of SARS-CoV-2 nucleocapsid antigen in 15 minutes (2). Individuals self-collect a nasal swab, place this in the buffer reservoir of the test cassette, close and seal the cassette, and read the results on a lateral flow strip.

The objective of this study was to store used Binax tests at ambient temperature with materials commonly found in the home and evaluate the performance of SARS-CoV-2 rRT-PCR and viral genome sequencing following RNA extraction from the test cassettes.

## Materials and Methods

### Ethics statement

The study protocol was reviewed and approved by the Emory Institutional Review Board. Participants provided informed consent to collect samples and data specifically for this study, except for three individuals from whom residual, de-identified home Binax tests were obtained in the clinic. Demographic and clinical data were not available from these individuals.

### Analytical evaluation

Eight samples (6 self-collected samples and 2 contrived samples) were used to evaluate the quantity of SARS-CoV-2 RNA extracted from different parts of the Binax test. Used Binax cassettes were disassembled, the swab was removed, and the pad that contacts the swab was separated from the cassette. The swab and pad were each placed into 200µL of MagMAX Viral/Pathogen Binding Solution (Thermo Fisher Scientific, Waltham, MA).

After observing better RNA recovery from swabs, dilution series of pooled samples containing SARS-CoV-2 delta (n=11 dilutions) and omicron variant (sublineage BA.1, n=12 dilutions) were used to evaluate the analytical performance of the extraction protocol. 50µL of each dilution was spiked onto swabs provided with the Binax test in duplicate. The tests were completed according to manufacturer instructions. For one concentration of delta and omicron, swabs were retrieved immediately after test completion (day 0) or stored at room temperature in the Binax cassette for 2 and 7 days. For the remaining dilutions, swabs were retrieved on day 0.

### Clinical samples

From May to October 2022, self-administered Binax tests were obtained from 31 participants. Demographic and clinical data were recorded. Following test completion, Binax cassettes were stored at room temperature in zipper-locked bags with the desiccant packet provided within each Binax kit until nucleic acid extraction could be performed (Figure S1).

### Nucleic acid extraction and rRT-PCR

Binax swabs were swirled in the tube containing Binding Solution 10 times and removed while squeezing out buffer against the side of the tube. Sample pads were incubated for 10 minutes. For all samples, the entire volume of Binding Solution was used as the starting material for extraction in the MagMAX Viral/Pathogen Nucleic Acid Isolation Kit with elution into 50µL of Tris-HCl, performed on a KingFisher Apex instrument (Thermo Fisher Scientific) according to manufacturer recommendations. After extraction, eluates were tested with three rRT-PCRs: 1) a multiplex reaction for Flu A, Flu B, the N2 target and RNase P (Flu-SC2) (11); 2) Spike SNP with 5 probes for K417, L452R, T478K, E484K, N501Y (5, 6); and 3) a triplex assay for *spike* Δ69/70 and ORF1a Δ3675-3677 (7). Assays were performed as previously described except for Flu-SC2, modified as shown in Table S1.

### Sequencing

Extracted RNA was used to synthesize cDNA using SuperScript IV (Invitrogen), and libraries were constructed using the xGen SARS-CoV-2 Amplicon panel (IDT) following the manufacturer’s protocol. Libraries were quantified using KAPA universal complete kit (Roche), pooled to 4nM and sequenced on an Illumina Miseq with paired-end 150-bp reads. Consensus viral genome sequences were assembled using viralrecon v2.4.4 (12). Pangolin v1.16 was used for lineage assignment (13).

### Phylogenetic analysis

Forty-five sequences obtained from 19 participants were aligned with reference sequences Wuhan/Hu-1/2019 and Wuhan/WHO/2019, as well as 266 SARS-CoV-2 sequences collected in Georgia between 20 May and 06 October 2022 (14). One sequence from participant 7, a traveler returning from Kenya, was aligned with 2,796 BA.5.2.1 sequences collected between 29 May and 06 June 2022. Reference sequences were downloaded from the Global Initiative on Sharing Avian Influenza Data (GISAID) (14). Alignments were performed using Nextalign within the Nextstrain v3.2.4 pipeline (15). For ease of visualization, the Georgia dataset was downsampled to 100 sequences using Nextstrain subsampling scheme “all”. The BA.5.2.1 dataset was downsampled to 50 sequences using a custom scheme, in which crowd penalty was set to 0.0 and proximity filter was set to the traveler sequence to select the most genetically related sequences. Maximum likelihood phylogenetic trees were constructed using default settings of the Nextstrain SARS-CoV-2 Workflow with TreeTime v0.8.6 8. Trees were visualized using Auspice v2.37.3.

### Statistical analysis

Calculation of means and standard deviations were done in Excel software (IBM). ANOVA and two-sided t-tests were performed in GraphPad Prism, version 9.3.1 (GraphPad Software).

## Results

### Analytical evaluation

For eight samples, RNA was extracted from both the swab and specimen pad in the Binax cassette. Ct values were significantly lower (higher RNA concentration) from the swabs (Figure S2), which were then used in all subsequent evaluations. Compared to the original Cts for delta and omicron dilutions, Cts for RNA extracted from Binax swabs on day 0 were a mean of 7.4 (standard deviation, 1.9) and 7.3 (1.1) cycles later, respectively (Table S2). All samples with a detectable test line on the Binax card were positive in the Flu-SC2 (Figure S3). In addition, the two omicron dilutions with no visible test line had detectable SARS-CoV-2 RNA. SARS-CoV-2 RNA remained detectable for at least 7 days of storage on the swab at room temperature, though Ct values increased with time (Figure S2B).

### Clinical Evaluation

103 Binax samples were obtained from 31 participants. Demographic and sample data are shown in Table S3. Binax test results were positive for 89/103 samples (86.4%). Following extraction from the used swab, 81/103 samples (78.6%) yielded detectable SARS-CoV-2 RNA in the Flu-SC2 assay (Table S4). SARS-CoV-2 RNA was detectable in six swabs despite negative Binax results, including two individuals with longitudinal sampling, who were symptomatic and RNA positive prior to Binax antigen detection. All samples were positive for RNase P detection, demonstrating successful nucleic acid extraction and absence of PCR inhibitors. Mean duration of Binax swab storage was 4.6 days (SD 3.2), and SARS-CoV-2 RNA remained detectable following swab storage for up to 16 days prior to extraction. The duration of storage was not significantly different for samples that tested positive in the Flu-SC2 assay versus negative (Figure S4). In rRT-PCRs to detect specific SARS-CoV-2 mutations, 66/81 Flu-SC2-positive samples (81.5%) tested positive in the Spike SNP assay and 74/81 samples (91.4 %) tested positive in the triplex assay for *spike* Δ69/70 and ORF1a 3675-3677. Results from Spike SNP and triplex testing were all consistent with omicron sublineage BA.2, BA.4, or BA.5.

### Sequencing

Full viral genome sequencing was attempted from 57 samples with Ct values less than 32: 46 samples from 23 participants yielded complete, high-quality sequences (Table S5). Cts of unsuccessful samples were high (range 29.0-34.3, mean-32.0) compared to successfully sequenced samples (range 15.6-33.7, mean-26.2). Pangolin lineage assignments confirmed Spike SNP and triplex assay results (Table S5).

### Specific applications of SARS-CoV-2 sequencing from Binax swabs

#### Longitudinal sampling

Time courses from eight participants who provided sequential samples during their illness demonstrated consistent and concordant SARS-CoV-2 detection by the Binax test and rRT-PCR from the swab throughout most of the symptomatic period (Figure 1 and Figure S5). However, this was flanked by periods of inconsistent test results during the first 1-2 days of symptoms and again as symptoms waned. Interestingly, in several individuals, rRT-PCR turned positive prior to Binax antigen results (Figures 1 and S5). One individual self-tested twice-a-day; no consistent difference in Flu-SC2 Ct value was observed between morning and evening (Figure 1D).

**Figure 1.**
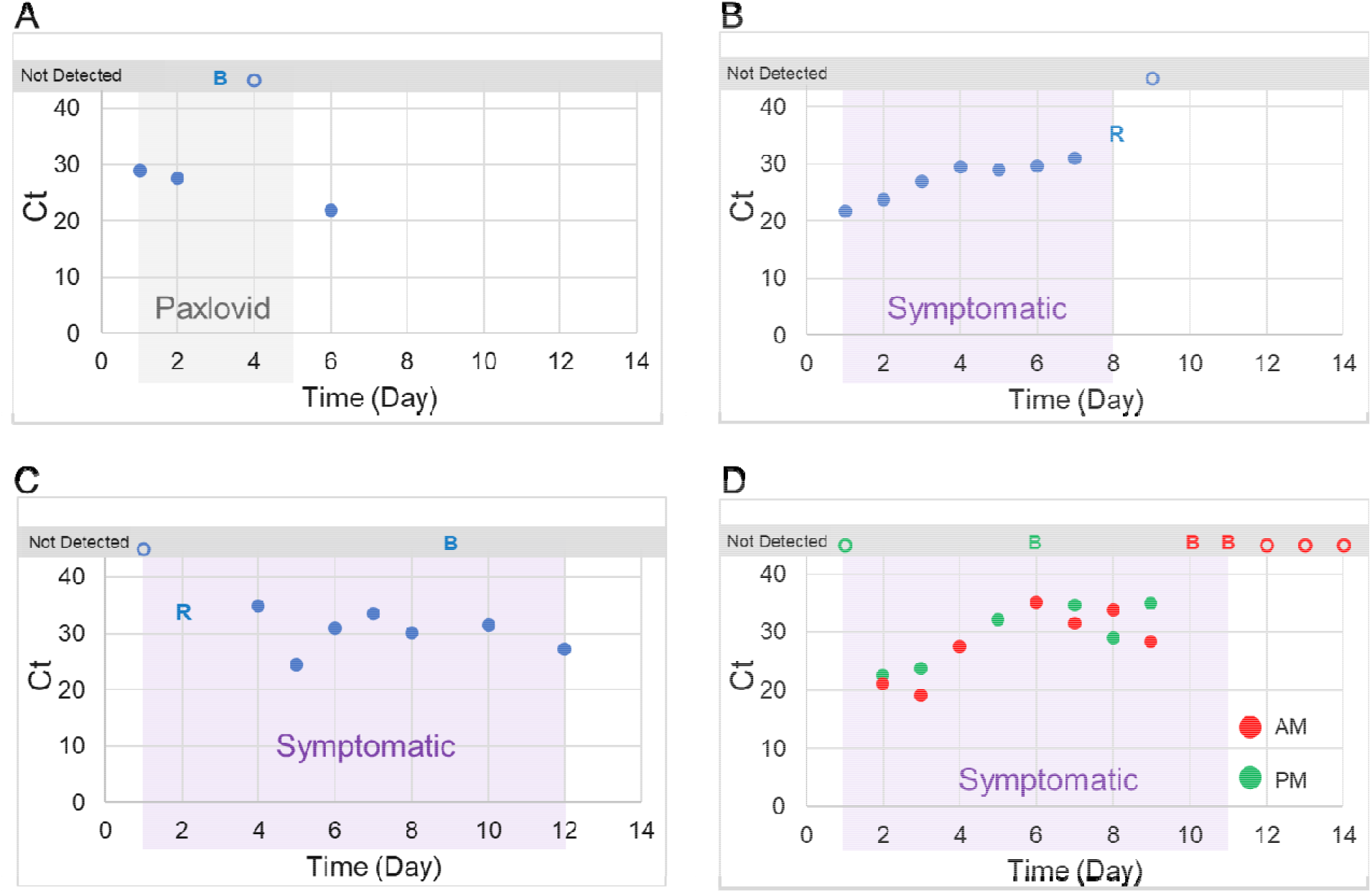
**A-D**) Time course of Binax results and SARS-CoV-2 rRT-PCR Cts using RNA extracted from Binax cassettes versus days post-symptom onset for four participants in the clinical evaluation. A) Gray shaded area indicates time on treatment with Paxlovid. B-D) Purple shaded areas indicate the symptomatic period. The participant for time course D collected AM and PM samples for 8 days. Symbols show the following Binax and rRT-PCR (Flu-SC2) results for individual samples: closed circles, both positive; B, Binax positive only; R, rRT-PCR positive only; open circle, both negative.

Phylogenetic analysis revealed that sequential samples from seven participants clustered by participant (Figure 2A). Within each participant, sequences were either identical or contained 1-2 SNPs relative to the earliest sequence. Sequences from three participants (1, 2, and 3), who were known close contacts, were all identical (Figure 2A).

**Figure 2.**
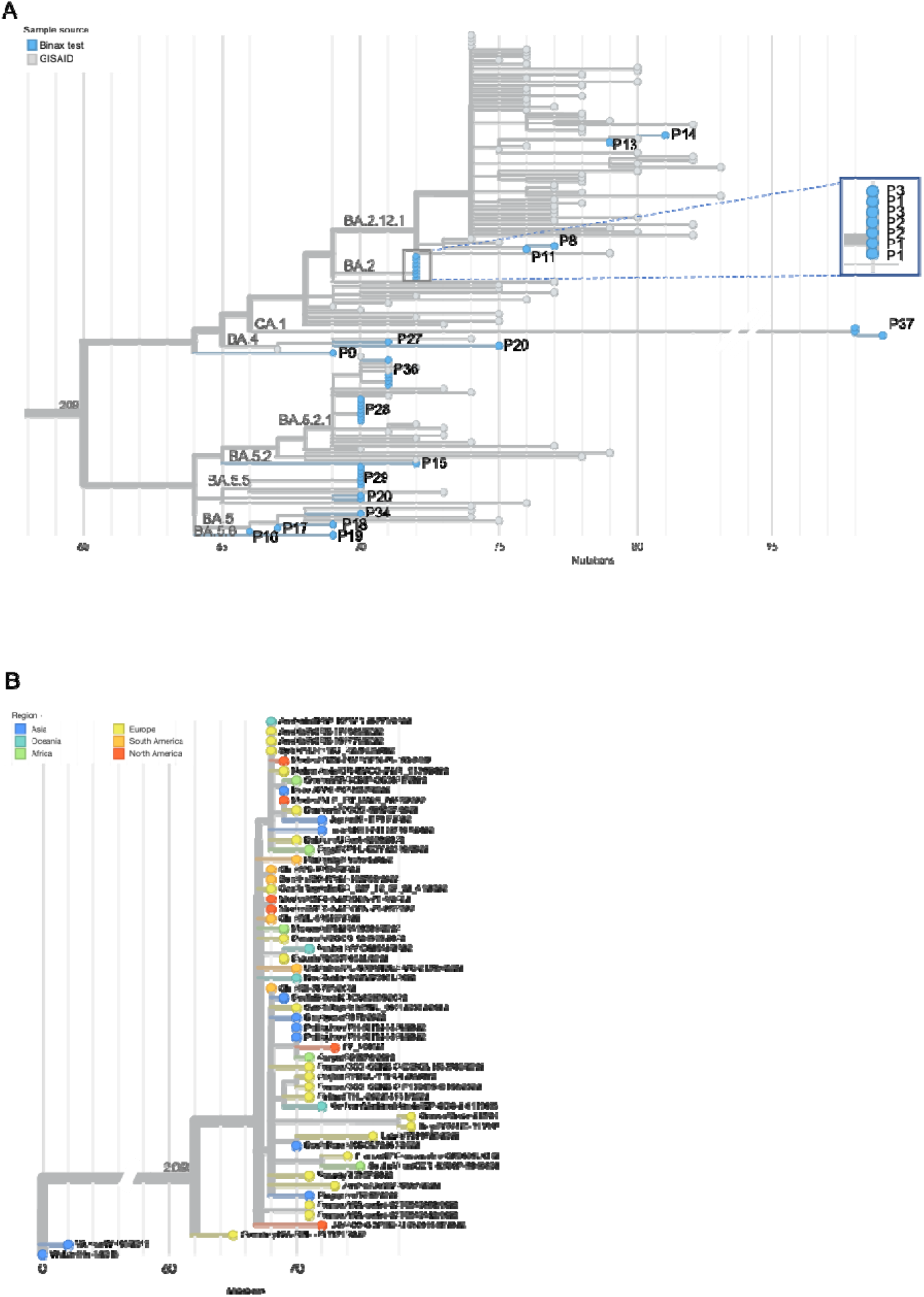
**A-B)** SARS-CoV-2 phylogenetic analysis of RNA extracted from used Binax tests. A) Phylogenetic tree contains SARS-CoV-2 sequences from 20 participants and illustrates: clustering of sequences from participants P1-P3, who had known epidemiologic linkage; clustering of longitudinal sequences in participants P28, P29, P36, and P37; and potential reinfection in participant P20. B) Phylogenetic tree illustrates clustering of sequence from participant P7, a returning traveler, with international sequences.

#### Antiviral treatment

One participant (20) with longitudinal sampling received Paxlovid, and Binax results and SARS-CoV-2 RNA converted from positive to negative on day 3 of antiviral treatment (Figure 1A). However, the day following discontinuation, the participant experienced a return of symptoms and a rebound in both Binax and RNA positivity. The sequence from this last sample did not cluster with the others and was a different lineage, indicating possible reinfection (Figure 2A).

#### Returned traveler

Participant 7 developed COVID-19 following travel to Kenya. Sequencing identified omicron sublineage BA.5.2.1, and phylogenetic analysis revealed that the sequence was closely related to a sequence from Kenya, supporting a travel related infection (Figure 2B).

## Discussion

This study demonstrates that SARS-CoV-2 RNA, extracted from used Binax test swabs, is of sufficient quality for genotyping by rRT-PCR and whole genome sequencing for phylogenetic studies. With a storage protocol that was purposefully kept simple to improve acceptability and feasibility, SARS-CoV-2 RNA remained detectable for up to 16 days. These data add to the scant literature on viral RNA recovery from rapid diagnostic tests (16-18), including for SARS-CoV-2 (19, 20), and importantly extend available information to include the Binax test, which is a common self-test sold directly to consumers in the United States (2). Wide implementation of SARS-CoV-2 sequencing from home tests could substantially enhance population-level surveillance of emerging variants, including from populations who may be otherwise under-represented among current surveillance approaches (9, 10, 21).

Furthermore, this study describes a set of use cases for the protocol beyond surveillance. Time courses demonstrate that sequential samples can be conveniently collected for the study of viral dynamics and monitoring within-host virus evolution. This may be particularly useful for identifying mutations in immunocompromised patients or following antiviral treatment (22). Notably, we identified a rebound in SARS-CoV-2 viremia following Paxlovid that may have resulted from re-infection. Additionally, phylogenetic clustering of sequences from known contacts demonstrates the use of this method to evaluate potential transmission events. Finally, we demonstrate the feasibility of using this method to detect potential SARS-CoV-2 introductions following international travel.

A limitation of this study was the lack of concurrent standard-of-care molecular testing. However, this would not have been feasible for all participants across all time points. Ct results may have improved with a shorter, defined duration of Binax storage, but this would have impacted convenience for participants and feasibility for laboratory testing.

In conclusion, SARS-CoV-2 RNA extraction from used Binax swabs stored at ambient temperature combines the convenience of rapid diagnostic results with the potential for genomic surveillance from home. This approach greatly facilitates investigation into viral dynamics, transmission clusters, and intra-host viral evolution.

## Data Availability

All sequences newly generated in this study were deposited in NCBI under BioProject PRJNA634356 and GISAID under accession numbers EPI_ISL_15938303-331. Sequences downloaded from GISAID are available at doi: 10.55876/gis8.221128zq.

## Acknowledgements

We gratefully acknowledge all data contributors, i.e., the Authors and their Originating laboratories responsible for obtaining the specimens, and their Submitting laboratories for generating the genetic sequence and metadata and sharing via the GISAID Initiative, on which this research is based.

## Author Contributions

Conceptualization, P-V. N., L.R.C., J.J.W., A.P.; Methodology, P-V. N., L.R.C., L.B., A.R., J.J.W., A.P.; Software, P-V. N., L.R.C., J.J.W., A.P.; Validation, P-V. N., L.R.C., L.B., A.R., J.J.W., A.P.; Formal Analysis, P-V. N., L.R.C., J.J.W., A.P.; Investigation, P-V.N., L.R.C., E.W., L.B., A.R., J.J.W., A.P.; Resources, P-V.N., L.R.C., L.B., A.R., M.G., J.A.S., G.S.M., W.A.L, J.J.W., A.P.; Data Curation, P-V. N., L.R.C., J.J.W., A.P.; Writing – Original Draft Preparation, P-V.N., L.R.C., J.J.W., A.P.; Writing – Review & Editing, P-V.N., L.R.C., E.W., L.B., A.R., M.G., J.A.S., G.S.M., W.A.L, J.J.W., A.P.; Visualization, P-V. N., L.R.C., M.G., W.A.L., J.J.W., A.P.; Supervision, J.J.W., A.P.; Project Administration, M.G., J.A.S., J.J.W., A.P.; Funding Acquisition, J.A.S, G.S.M, W.A.L.

## Institutional Review Board Statement

The study protocol was reviewed and approved by the Emory Institutional Review Board.

## Informed Consent Statement

Participants provided informed consent to collect samples and data specifically for this study, except for three individuals from whom residual, de-identified home Binax tests were obtained in the clinic. Demographic and clinical data were not available from these individuals.

## Data Availability

All sequences newly generated in this study were deposited in NCBI under BioProject PRJNA634356 and GISAID under accession numbers EPI_ISL_15938303-331. Data availability of sequences downloaded from GISAID is listed in Table S6.

