## Supplemental Figures and Tables for "SARS-CoV-2 molecular testing and whole genome sequencing following RNA recovery from used BinaxNOW COVID-19 Antigen Self Tests"

**Supplemental Material**

Phuong-Vi Nguyen and Ludy Registre Carmola, Ethan Wang, Leda Bassit, Anuradha Rao, Morgan Greenleaf, Julie A. Sullivan, Greg S. Martin, Wilbur A. Lam, Jesse J. Waggoner and Anne Piantadosi

**Supplemental Figures**

**Figure S1.** Workflow of patient Binax sample collection, storage and shipment at ambient temperatures, RNA extraction and molecular testing. Created with BioRender.com.

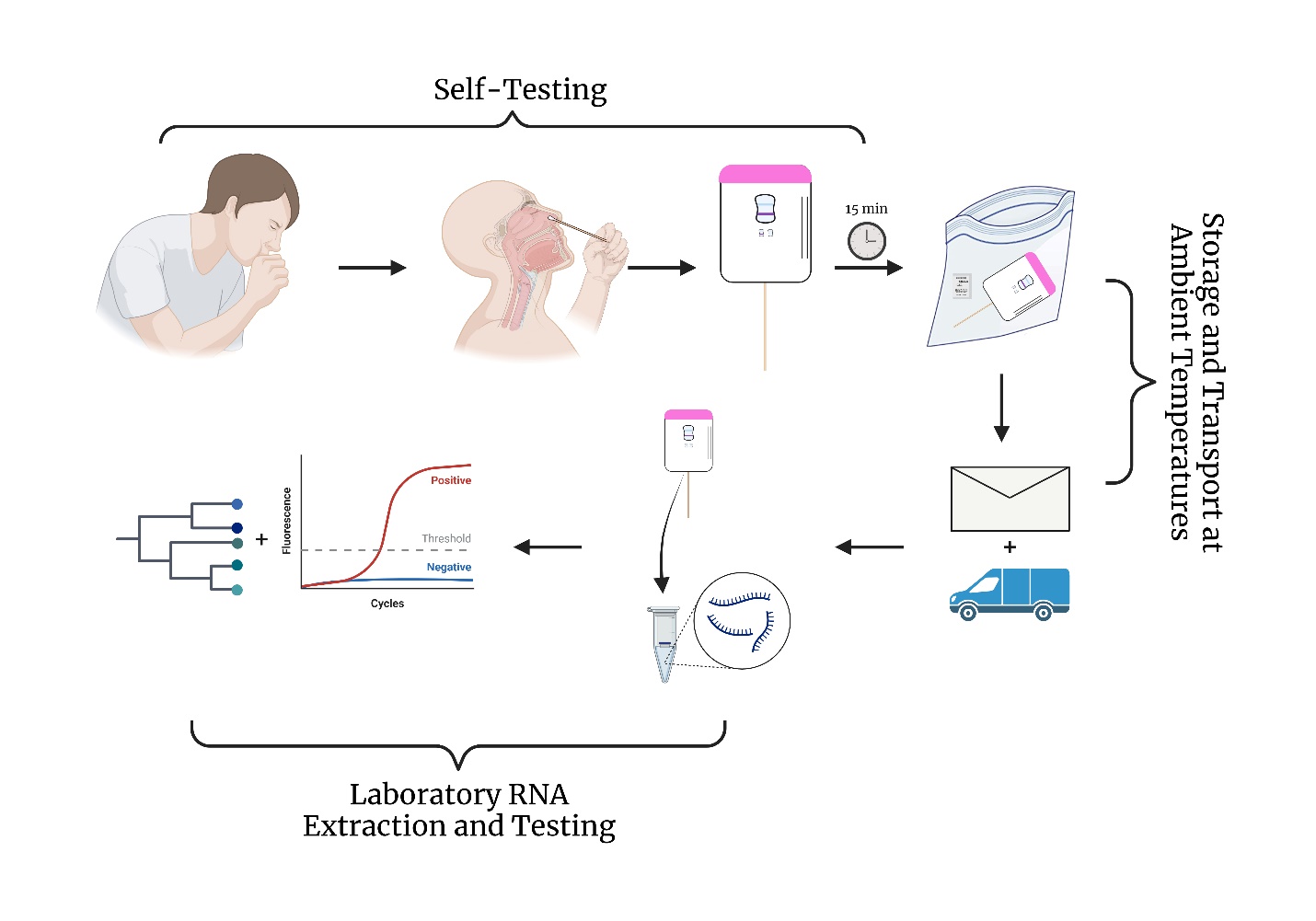

**Figure S2. A**) Comparison of SARS-CoV-2 Ct in the Flu-SC2 assay following nucleic acid extraction from used Binax swabs or the specimen pad from the cassette. **B**) Stability of SARS-CoV-2 RNA stored in the Binax test cassette at room temperature for 0, 2, and 7 days.

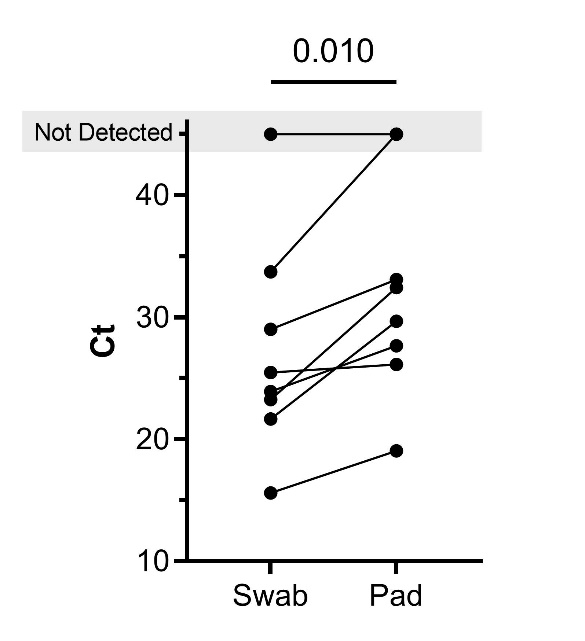

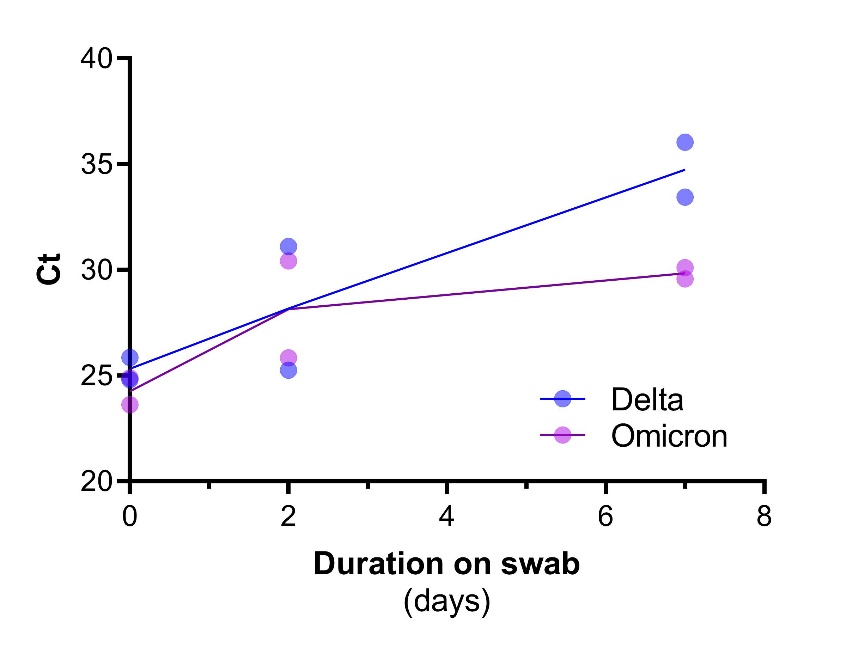

**A**

**B**

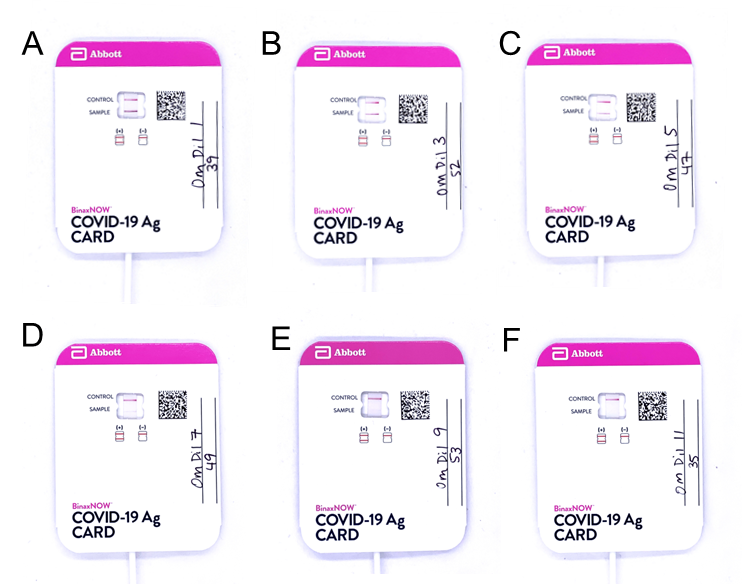
**Figure S3.** Images of positive (**A-E**) and negative (**F**) Binax tests using Omicron dilutions from which SARS-CoV-2 RNA was successfully extracted from the swab and detected by rRT-PCR. Binax cassettes displayed correspond to Omicron dilutions 1, 3, 5, 7, 9, and 11 in Table S2.

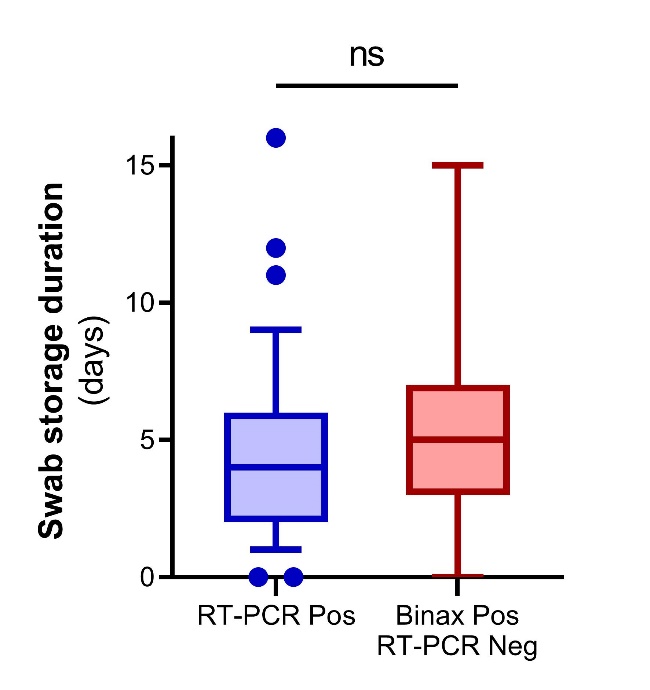
**Figure S4.** Duration of swab storage in used Binax cassettes was not significantly different for samples positive by rRT-PCR for SARS-CoV-2 (Flu-SC2) after RNA extraction from used swabs versus samples that were Binax positive/rRT-PCR negative. Box shows median and interquartile range; whiskers extend from 5-95% of all values.

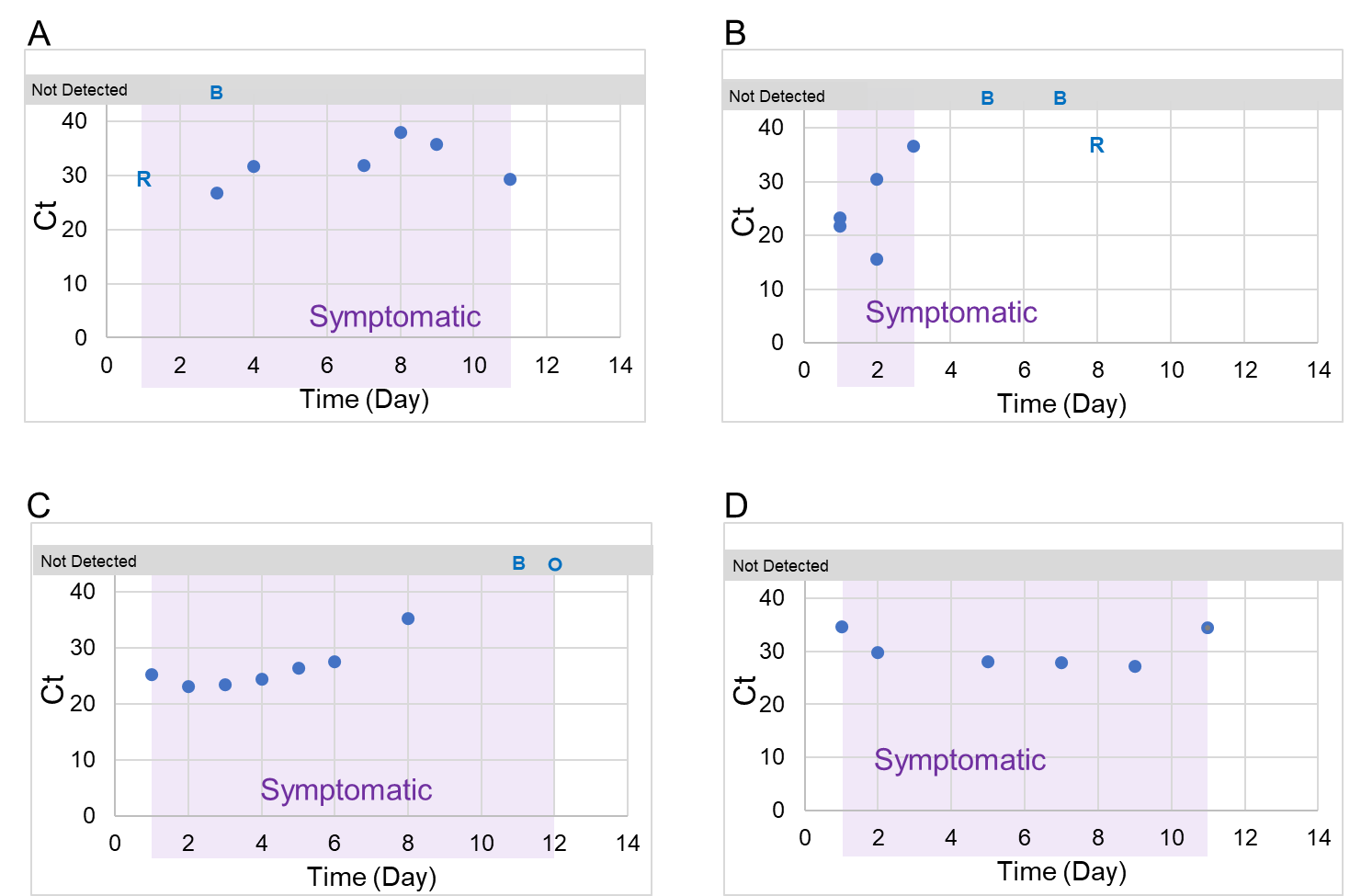

**Figure S5. (A-D)** Time-course of Binax results and SARS-CoV-2 rRT-PCR Cts using RNA extracted from Binax cassettes versus days post-symptom onset for four additional participants in the clinical evaluation. Purple shaded areas indicate the symptomatic period. Symbols show the following Binax and rRT-PCR (Flu-SC2) results for individual samples: closed circles, both positive; B, Binax positive only; R, rRT-PCR positive only; open circle, both negative.

**Table S1.** Flu-SC2 assay as performed in the current study.

| **Oligonucleotide** | **Sequence** | **Concentration ^a^** |
| --- | --- | --- |
| ***Primers*** |  |  |
| Influenza A Forward 1 | CAAGACCAATCYTGTCACCTCTGAC | 200nM |
| Influenza A Forward 2 | CAAGACCAATYCTGTCACCTYTGAC | 200nM |
| Influenza A Reverse 1 | GCATTYTGGACAAAVCGTCTACG | 200nM |
| Influenza A Reverse 2 | GCATTTTGGATAAAGCGTCTACG | 200nM |
| Influenza B Forward | TCCTCAAYTCACTCTTCGAGCG | 400nM |
| Influenza B Reverse | CGGTGCTCTTGACCAAATTGG | 400nM |
| SARS-CoV-2 Forward | CTGCAGATTTGGATGATTTCTCC | 400nM |
| SARS-CoV-2 Reverse | CCTTGTGTGGTCTGCATGAGTTTAG | 400nM |
| RNase P Forward | AGATTTGGACCTGCGAGCG | 100nM |
| RNase P Reverse | GAGCGGCTGTCTCCACAAGT | 100nM |
| ***Probes*** |  |  |
| Influenza A | FAM – TGCAGTCCTCGCTCACTGGGCACG – BHQ1 | 200nM |
| Influenza B | CF560 – CTGTGTTCATAGCTGAGACCATCTGC – BHQ1 | 200nM |
| SARS-CoV-2 | CF610 – ACAATTTGCCCCCAGCGCTTCAG – BHQ2 | 200nM |
| RNase P | Q705 – TTCTGACCTGAAGGCTCTGCGCG – BHQ2 | 50nM |

Abbreviations: BHQ1/2, Black Hole Quencher 1/2; CF560, Cal Fluor Orange 560; CF610, Cal Fluor Red 610; FAM, fluorescein; Q705, Quasar 705

^a^ Concentration in the final 20µL reaction mixture of the Luna Universal One-Step RT-qPCR Kit (New England Biolabs). Reactions were performed with the following cycling conditions: 52°C for 15 min; 94°C for 2 min; and 45 cycles at 94°C for 15s, 55°C for 40s (signal acquired), 68°C for 20s.

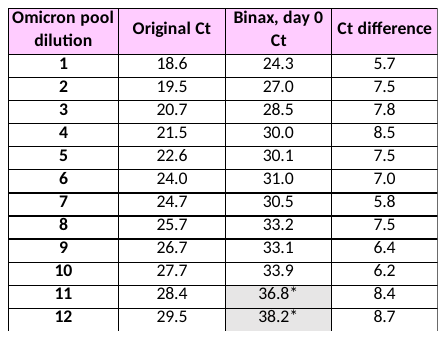

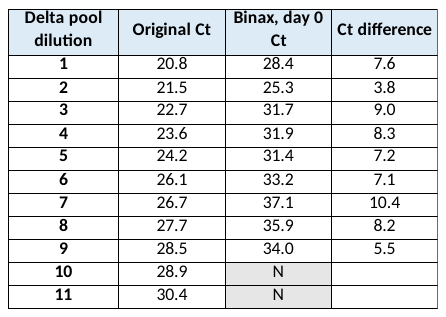
**Table S2.** Cts for the original dilution series of heat-inactivated, pooled samples of delta and omicron (BA.1) variants compared to Cts following RNA extraction from used Binax swabs. Shaded boxes indicate negative Binax results.

**Table S3.** Characteristics of participants in the clinical evaluation.

| **Characteristic** | **Participants** |
| --- | --- |
| Total, n | 31 |
| Age, years, mean (SD) | 34.9 (18.3) |
| Gender, female, n (%) | 17 (54.8) |
| Vaccinated, n (%) | 24 (77.4) |
| Time since last dose, days, mean (SD)^a,^ | 180.8 (109.4) |
| Symptomatic, yes, n (%) | 30 (96.8) |
| Samples, n | 103 |
| Symptomatic at collection, yes, n (%) | 89 (86.4) |
| Time on swab, days, mean (SD)^b^ | 4.6 (3.2) |

Abbreviations: SD, standard deviation

Missing data: 2 participants for age, gender, vaccination status, and time since last dose; 2 samples for symptomatic at collection

^a^ Defined as time from last vaccine dose to first swab collected.

^b^ Defined as time from swab collection to RNA extraction.

**Table S4.** Comparison of the called result from the BinaxNOW COVID-19 Antigen Self Test and SARS-CoV-2 rRT-PCR following RNA extraction from used swabs. SARS-CoV-2 rRT-PCR positive was defined as RNA detection in the modified Flu-SC2 assay.

|  |  | **Binax Call** | | |
| --- | --- | --- | --- | --- |
|  |  | **Positive** | **Negative** | **Total** |
| **SARS-CoV-2 rRT-PCR** | **Positive** | 75 | 6 | 81 |
|  | **Negative** | 14 | 8 | 22 |
|  | **Total** | 89 | 14 | 103 |

**Table S5.** Whole genome sequencing metrics (coverage, depth, etc.) of Binax swab samples collected May–October 2022.

| **Sample** | **Ct Value** | **Mapped reads** | **Coverage (%)** | **Depth** | **Length** | **Lineage** |
| --- | --- | --- | --- | --- | --- | --- |
| P1-4704X | 29.0 | 276,350 | 96 | 427 | 29,873 | BA.2 |
| P1-4712F | 26.7 | 283,368 | 99 | 826 | 29,873 | BA.2 |
| P1-5650R | 29.3 | 924,498 | 99 | 1,648 | 29,873 | BA.2 |
| P2-4706Z | 23.2 | 1,006,030 | 100 | 3,081 | 29,873 | BA.2 |
| P2-4709C | 15.6 | 1,462,734 | 100 | 2,317 | 29,873 | BA.2 |
| P2-4705Y | 21.7 | 1,442,693 | 99 | 3,180 | 29,873 | BA.2 |
| P3-4707A | 33.7 | 78,193 | 73 | 87 | 29,882 | BA.2 |
| P3-4715I | 24.6 | 153,717 | 96 | 280 | 29,873 | BA.2 |
| P3-5636D | 27.3 | 1,804,990 | 99 | 3,388 | 29,873 | BA.2 |
| P5-4902N | 29.8 | 228,743 | 99 | 674 | 29,876 | BA.2-like |
| P7-4901M | 21.8 | 414,525 | 99 | 1161 | 29,870 | BA.5 |
| P8-5628V | 23.9 | 402,061 | 99 | 987 | 29,876 | BA.2.12.1 |
| P9-5635C | 26.7 | 629,820 | 100 | 2,098 | 29,876 | BA.2 |
| P10-5633A | 25.7 | 327,250 | 99 | 830 | 29,876 | BA.5 |
| P11-5640H | 25.6 | 1,718,284 | 100 | 4,289 | 29,876 | BA.2.12.1 |
| P13-5646N | 28.4 | 588,636 | 99 | 1,059 | 29,876 | BA.2.12.1 |
| P14-5639G | 27.5 | 635,555 | 98 | 946 | 29,885 | BA.2.12.1 |
| P15-5642J | 27.7 | 342,821 | 96 | 508 | 29,870 | BA.5.2 |
| P17-5621O | 21.2 | 1,583,935 | 99 | 4,073 | 29,870 | BA.5.1 |
| P18-5643K | 28.2 | 417,631 | 97 | 621 | 29,870 | BA.5.1 |
| P19-5632Z | 25.4 | 232,062 | 96 | 334 | 29,870 | BA.5.6 |
| P20-5622P | 21.9 | 1,954,921 | 100 | 5,505 | 29,861 | BA.4 |
| P20-5638F | 27.5 | 689,409 | 98 | 1,344 | 29,870 | BA.5.5 |
| P20-5647O | 29.0 | 891,213 | 97 | 1,815 | 29,870 | BA.5.5 |
| P27-5644L | 28.2 | 1,588,267 | 99 | 3,496 | 29,861 | BA.4 |
| P28-5619Q | 19.1 | 2,764,958 | 99 | 6,851 | 29,879 | BA.5.2.1 |
| P28-5620R | 21.0 | 1,979,547 | 99 | 5,396 | 29,879 | BA.5.2.1 |
| P28-5623Q | 22.5 | 523,433 | 98 | 1,240 | 29,870 | BA.5.1 |
| P28-5627U | 23.6 | 761,370 | 99 | 2,058 | 29,879 | BA.5.2.1 |
| P28-5637E | 27.5 | 703,852 | 98 | 1381 | 29,870 | BA.5.2.1 |
| P28-5645M | 28.4 | 802,225 | 98 | 1,443 | 29,870 | BA.5.2.1 |
| P28-5649Q | 29.0 | 396,660 | 98 | 711 | 29,870 | BA.5.2.1 |
| P29-5624R | 23.1 | 1,037,60 | 98 | 2,327 | 29,870 | BA.5.5 |
| P29-5625S | 23.4 | 1,305,699 | 99 | 2,563 | 29,870 | BA.5.5 |
| P29-5629W | 29.0 | 629,778 | 98 | 1,584 | 29,870 | BA.5.5 |
| P29-5631Y | 25.3 | 660,795 | 99 | 1,804 | 29,870 | BA.5.5 |
| P29-5634B | 26.3 | 150,746 | 99 | 448 | 29,879 | BA.5.5 |
| P29-5641I | 27.6 | 586,112 | 98 | 958 | 29,870 | BA.5.5 |
| P34-5626T | 23.5 | 1,014,847 | 99 | 2,643 | 29,870 | BA.5.1 |
| P36-6013U | Unavailable | 949,042 | 99 | 3089 | 29,870 | BA.5.2.1 |
| P36-6014V | Unavailable | 723,386 | 99 | 2555 | 29,879 | BA.5.2.1 |
| P36-6015W | Unavailable | 568,952 | 99 | 2101 | 29,879 | BA.5.2.1 |
| P36-6030L | Unavailable | 269,584 | 97 | 555 | 29,870 | BA.5.2.1 |
| P36-6031M | Unavailable | 383,548 | 98 | 747 | 29,870 | BA.5.2.1 |
| P37-6323S | 27.9 | 1,256,274 | 99 | 3,212 | 29,876 | CA.1 |
| P37-6324T | 27.8 | 888,928 | 99 | 2,083 | 29,876 | CA.1 |
| P37-6325U | 27.2 | 1,438,107 | 99 | 3,864 | 29,876 | CA.1 |

**Table S6.** GISAID Data availability

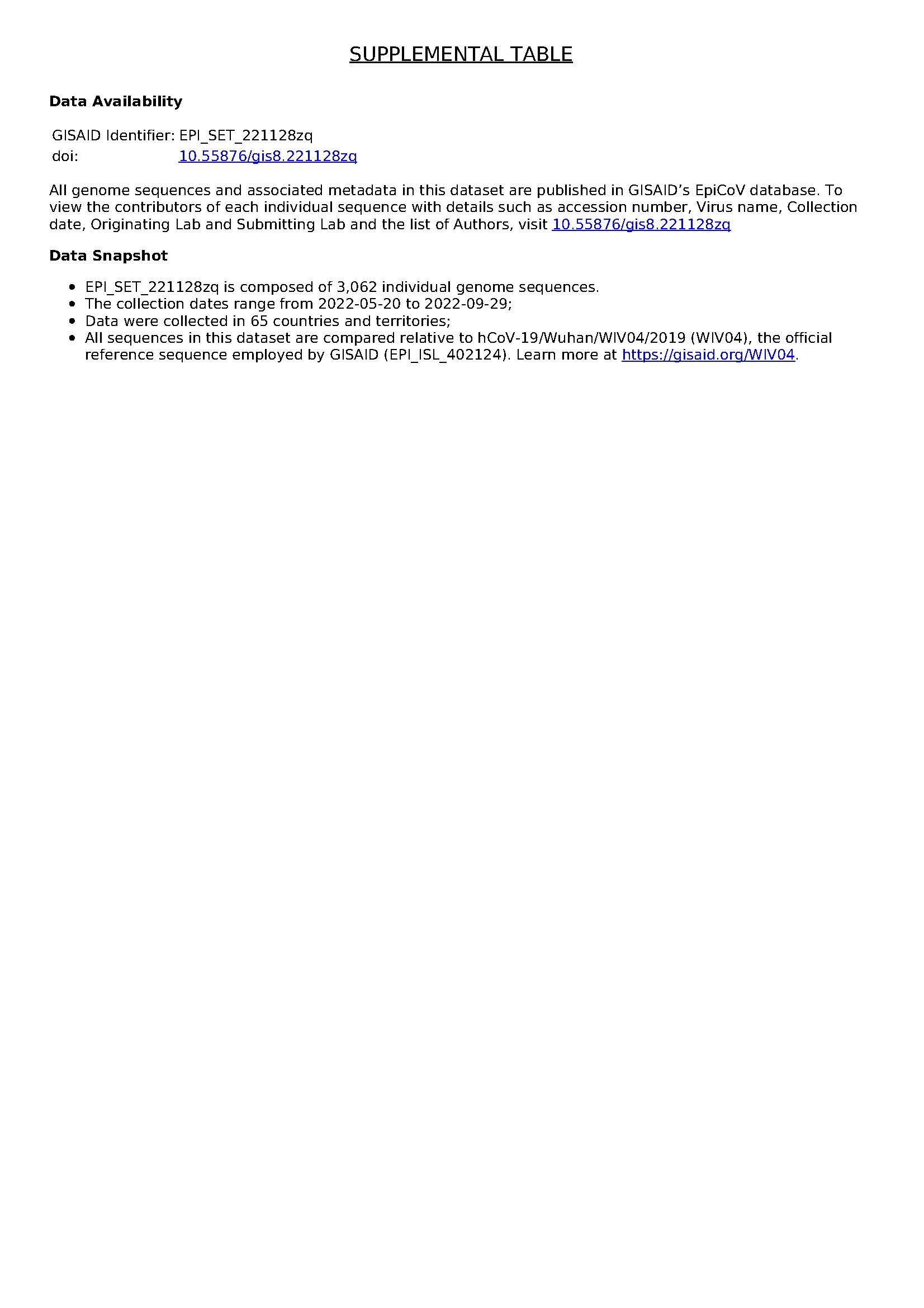
